# Experimental investigation to verify if excessive plastic sheeting shielding produce micro clusters of SARS-CoV-2

**DOI:** 10.1101/2021.05.22.21257321

**Authors:** Yo Ishigaki, Yuto Kawauchi, Shinji Yokogawa, Akira Saito, Hiroko Kitamura, Takashi Moritake

## Abstract

We experimentally investigated indoor air ventilation using the CO_2_ tracer technique to verify the infection cluster of SARS-CoV-2 that erupted at an office space. Multi-placed observations revealed extremely low air change rates (0.1/h) at the site. The local infection clusters were observed several meters away from a door that is the only ventilation in the office, which suggests a negative effect of plastic sheeting shielding. The thermo-fluid simulation showed that the plastic sheet blocked the airflow and trapped the exhaled air in each partition cell. As risk suppression methods, improving air ventilation by opening windows and using fans were verified, and significant improvements (10–28/h) were observed for each partition cells.

## Introduction

Coronavirus disease 2019 (SARS-CoV-2) has currently spread all over the world and has changed our lives profoundly^1)^. As it carries a high mortality rate, public precautions such as town lockdown and regulation of traffic and telework are considered. However, several workers are forced to gather in their office, and hence, offices and administrations should enhance ventilation and install small compartment partitions as recommended^2)^ by some local governments.

The infection route of SARS-CoV-2 is presented in the results and discussion section. The transmission of coronavirus 2 (SARS-CoV-2) through breath exhalation remains unclear, which can lead to severe acute respiratory syndrome. Contact and droplet infections are believed to contribute to the transmission of SARS-CoV-2, and maintaining social distancing will prevent transmission. Simultaneously, evidence for COVID-19 transmission via aerosols has been reported in the past few months ^3-9)^. The aerosol contains particles with diameters of less than 5 um that can float in the air for minutes to hours. Therefore, transmission is much easier and rapid indoors than outdoors.

Recently, a plastic sheeting shielding (PSS) that forms the small compartments is often being installed in offices to suppress the risk of infection. However, the architectonical effect of the compartments is still unclear as a preventive method for further spread of infection. The compartments will contribute to blocking aerosol transportation. Contrastingly, the concentration of the aerosol in a compartment where SARS-CoV-2 positive people stay will significantly increase and may cause a small-sized infection cluster.

In this study, we investigated the negative effect of the PSSs at the site of an infection cluster of COVID-19 using CO_2_ Tracer. In March 2021, an outbreak of COVID-19 in an office with PSSs in Miyagi, Japan, involved two small-sized and separated clusters. Air ventilations of each compartment and effects of PSSs were investigated by the CO_2_ tracer technique using dry-ice emission. Rise and fall patterns of the CO_2_ concentration of each compartment during an experiment were compared by time-series analysis technique. The effects of opening windows and using fans were experimentally verified to improve the ventilation of each compartment. Additionally, using thermo-fluid simulation to reproduce the situation at the time the clusters were generated, we confirmed that PSSs blocked the airflow and trapped the exhaled air in each partition cell.

## Methodology

In March 2021, an outbreak of COVID-19 affected 11 persons who worked at the same air-conditioned office room in Miyagi, Japan. The index case-patient experienced fever and cough and went to the hospital. Three others had become positive with COVID-19 in the following 3 days. After that, a total of seven people had become positive. The office is an air-conditioned three-floor building. The third-floor office area occupies 180 m^2^ (6 ⨯ 30 m^2^), and one side of the wall which faces the outside has windows. However, as the winter cold had set in, the windows were not opened to let in fresh air.

Some PSS had been installed in this office, and the indoor space was compartmentalized to five areas (A, B, C, D and E). Seven or eight desks had been arranged back-to-back in the compartments, as shown in Fig. 1. Area A is a dedicated space for the management level of this office. Patients had tested positive for the COVID-19 with the polymerase chain reaction (PCR) test. Many patients worked in area B and had tested positive with PCR test. However, despite working in area C next to area B, eight persons tested negative with the PCR test. The infection had spread to area D, and four out of eight persons tested positive.

**Fig. 1.**
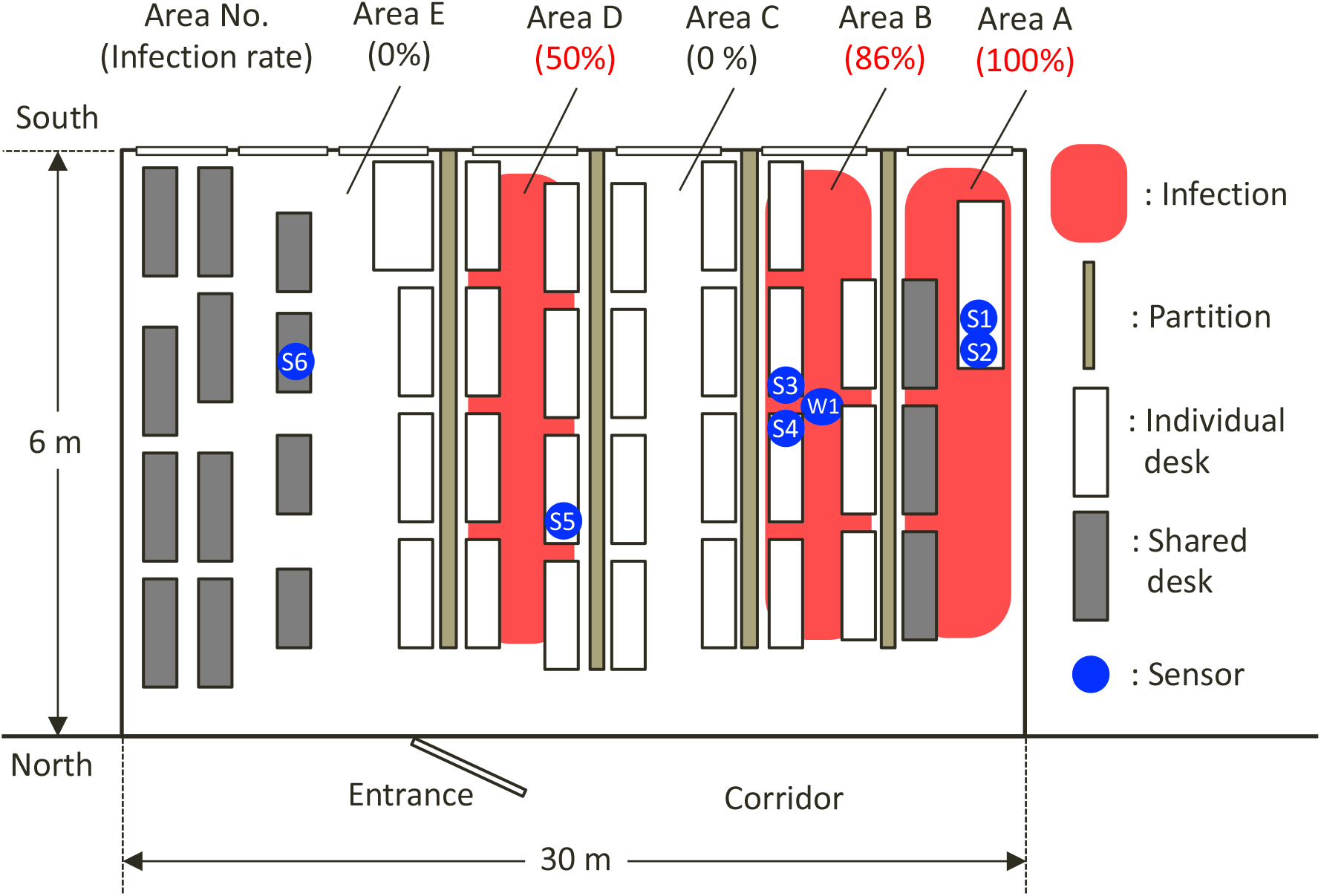
Synoptic view showing the arrangement of partitions and desks at the site of COVID-19 outbreak, Miyagi, Japan, 2021. Red-hatch zones indicate the area in that desks of case-patients were arranged; white zones indicate an unaffected desk area.

This skip infection spread for distant compartments gives two assumptions of the infection route for COVID-19. One is only the contact or droplet infections are dominant for COVID-19, which implies that aerosols do not affect the infection spread. However, some recent reports suggest that aerosol infection is one of the major routes of COVID-19 infection spread^3-9)^, and the potential of the first assumption is low. The other assumption is that the aerosol infection spread is prevented by the PSS to the outside of compartments but enhanced inside. The only known source of exposure for the affected persons in the office was the index case-patient. Additionally, all workers spent the same work hours with rules of hand disinfection and wearing masks for preventing transmissions in the office. These facts support the latter assumption and suggest a necessity of government guideline revision.

For the verification of the latter assumption, we investigated local indoor air ventilation experimentally. Indoor airflows were observed by using the CO_2_ tracer technique. As CO_2_ concentration can be accounted as an alternative to exhaled air amount, local infection risk could be evaluated indirectly. In this study, dry ice was used for CO_2_ emission, and several different types of CO_2_ sensors were used to detect the change of local CO_2_ concentration in the compartments. One wind sensor that can detect the direction and amount of wind was used to check the airflow in a compartment.

Two types of non-dispersive infrared (NDIR) gas sensors applied in an environmental field are used as CO_2_ sensors (Fig. 2). The TR-76Ui; T&D Corporation products, can detect CO_2_ concentration ranging from 0 to 9,999 ppm with ± 50 ppm ± 5% accuracy of measurement. The Pocket CO_2_ Sensor; YAGUCHI ELECTRIC CORPORATION products, can also detect CO_2_ concentration ranging from 400 to 10,000 ppm with ±30 ppm ±3% accuracy of measurement. Four TR-76Ui and two Pocket CO_2_ sensors were set up in the compartments as shown in Fig. 1. The symbols of S1, S3, S5, and S6 indicate TR-76Ui, and S2 and S4 indicate Pocket CO_2_ sensor. We developed our own wind sensor by combining a unidirectional airflow sensor OMRON D6F-W with a stepping motor. The wind sensor operates minutely while spinning in 360 degrees. The maximum wind volume and its direction are recorded. It was set up in the compartment of the index case-patient. The data from each sensor were collected in the database as soon as it was measured, which resulted in some disadvantages, such as the lack of periodicity and the necessity for data completion. In this study, we used PI System™ (OSIsoft) for data management, that is, to prepare the acquired data and convert it into tidy data^10)^ in which each variable is a column, each observation is a row, and each type of observational unit is a table with time synchronization.

**Fig. 2.**
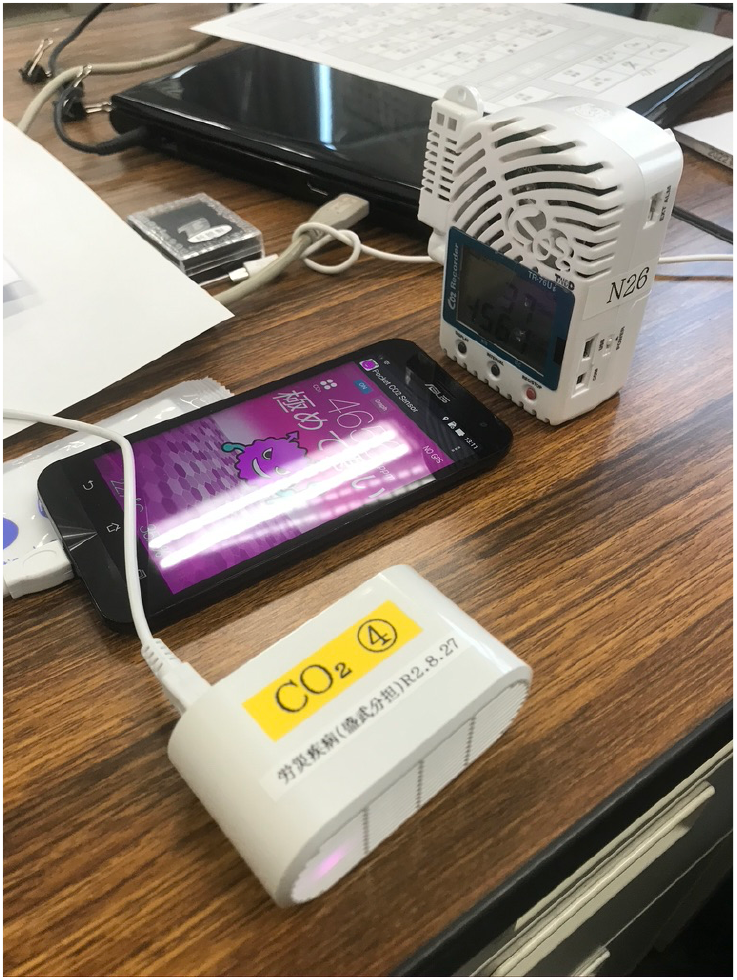
Photograph showing sensors used in this study. Upper: TR-76Ui; Lower: Pocket CO_2_ sensor.

We use CO_2_ not only as a risk proxy but also as a ventilation tracer. For the purpose of estimating the probability of airborne transmission of an infectious agent indoors the equation called the Wells–Riley model represented by the following equation^11)^ is widely used.

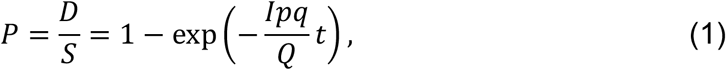

where *P* is the probability of infection, *D* is the number of disease cases, *S* is the number of susceptible people, *I* is the number of infectors, *p* is the breathing rate per person [m^3^/h], *q* is the quantum generation rate by an infected person [quanta/s], *Q* is the outdoor air supply [m^3^/h], and t is the total exposure time [s]. This model and its analogies are currently applied to evaluate COVID-19 infections. Especially, *Q* is a key factor for the infection risk of indoor environments. It suggests that the degree of inhomogeneity in indoor air is directly proportional to the risk distribution in the room. Recently, some researchers claimed that the exhaled CO_2_ can be treated as a COVID-19 infection risk proxy ^9, 12, 13^).

The local infection risk can be estimated by observing time-series change in CO_2_ concentration. Equation (2) derived from Seidel describes the concentration contaminant in a room ^14, 15^):

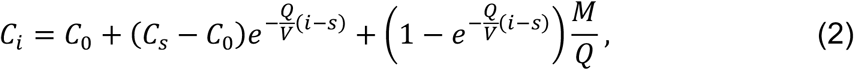

where *C*_*i*_ is a concentration of indoor pollutants at the time *i, C_0_* is a concentration in the absence of pollutant sources, *V* is a volume of the room, *s* is the time when CO_2_ emission is stopped, and *M* is the number of pollutants generated. In the state of no generation of pollutants, eq. (2) can be transforms to:

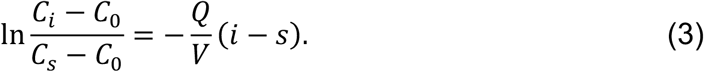

This equation suggests that a decrease in the normalized concentration of pollutants with ventilation time (i – s) under no generation corresponds to air change rate (*Q/V)* in the space as one of the pollutants, CO_2_, can be measured using sensors.

## Results and Discussion

Indoor CO_2_ concentration was increased by scattering crumbled dry ice on the floor without ventilation, as shown in Fig. 3. After the CO_2_ concentration at each point reached over 6000 ppm, the decrease in values were measured with three ventilation conditions as shown in Table 1. Condition 1 simulated the ventilation condition when the outbreak of COVID-19 was confirmed. Condition 2 investigates the effects of opening the windows on ventilation, and condition 3 investigates the effect of added fans on the ventilation at the windows to eject indoor air to the outside. Another condition was evaluated after a CO_2_ concentration decreased under 1000 ppm to suppress the effects of the previous condition.

**Table 1.** Experimental conditions.

|  | Condition 1 | Condition 2 | Condition 3 |
| --- | --- | --- | --- |
| Purpose | Replication of infections | Effects of window opening | Effects of window opening and fans |
| Room entrance | Open | Open | Open |
| Windows of corridor | Closed | Open | Open |
| Room windows | Closed | Open | Open |

| Fans | w/o fan | w/o fan | with fan |
| --- | --- | --- | --- |

**Fig. 3.**
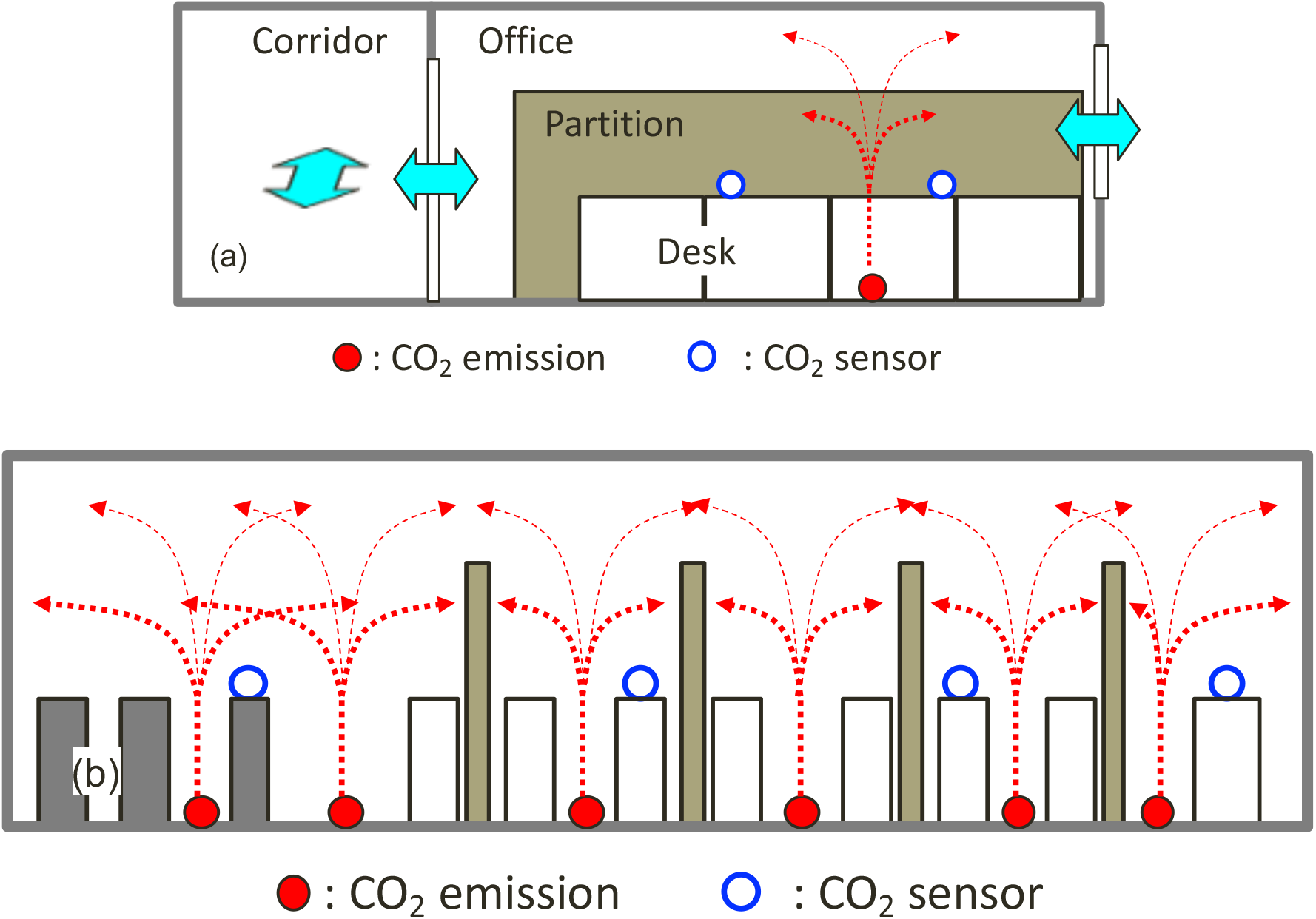
Cross-sectional conceptual diagram of experiments. Red arrows show CO_2_ emission and diffusion. Blue arrows indicate the ventilation of this office.

Figure 4 indicates a rise and fall in the CO_2_ concentration during an experiment. Concentration changes in the same compartments behaved in unison as compared to those of the other compartments. Additionally, CO_2_ concentrations reached different levels for each compartment, for both CO_2_ increasing and decreasing periods. The results obtained using the two types of sensors were in good agreement with these results. Temporal coincidence and similarity of pattern for the time-series data pairs were evaluated using two statistical indexes. The correlation coefficient indicates the temporal coincidence of time-series data from each sensor. The minimum cumulative distance calculated by the dynamic time warping method^16, 17^) (*L*_dtw_) indicates the similarity of CO_2_ change pattern that depends on ventilations around sensors. The calculated result matrix is shown in Fig. 5.

**Fig. 4.**
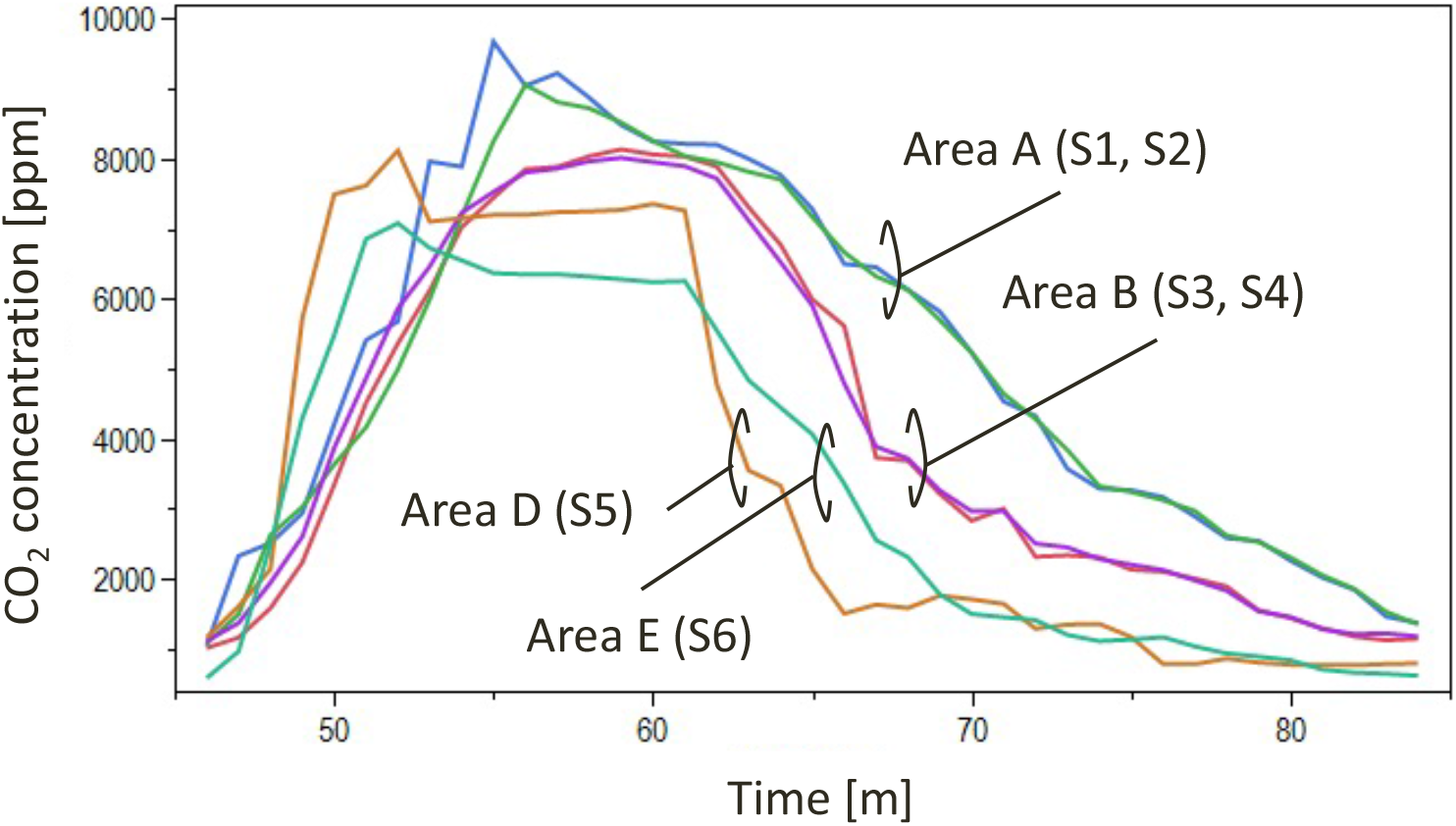
Line plot representing the CO_2_ concentration change in the experiment of condition 2.

**Fig. 5.**
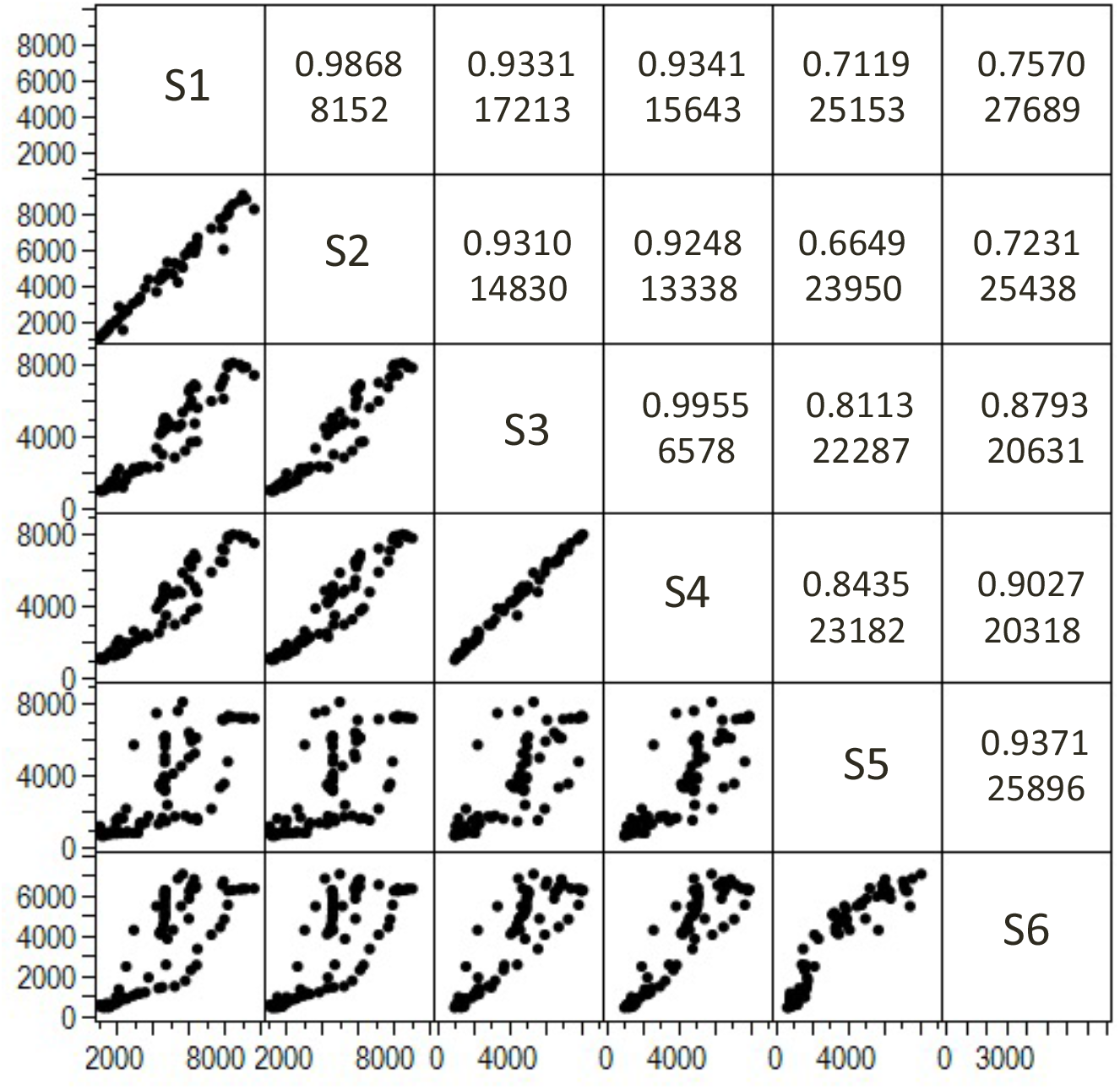
Correlation coefficient and minimum cumulative distance matrix of each sensor data pair.

Temporal coincidence and similarity of pattern is discussed on a scatter chart in Fig. 6. Sensors that were installed in the same compartment (S1 and S2, S2 and S4) showed a high correlation coefficient (>0.95) and comparatively small *L*_dtw_ Contrarily, sensors that were installed in the two adjacent compartments detected a larger *L*_dtw_ due to the difference in the CO_2_ change pattern, despite the high correlation coefficient (>0.90). More distant compartment pairs indicate a larger *L*_dtw_ irrespective of a correlation coefficient. These results suggest that the ventilation of the compartment is nearly independent due to the partition effects of PSSs.

**Fig. 6.**
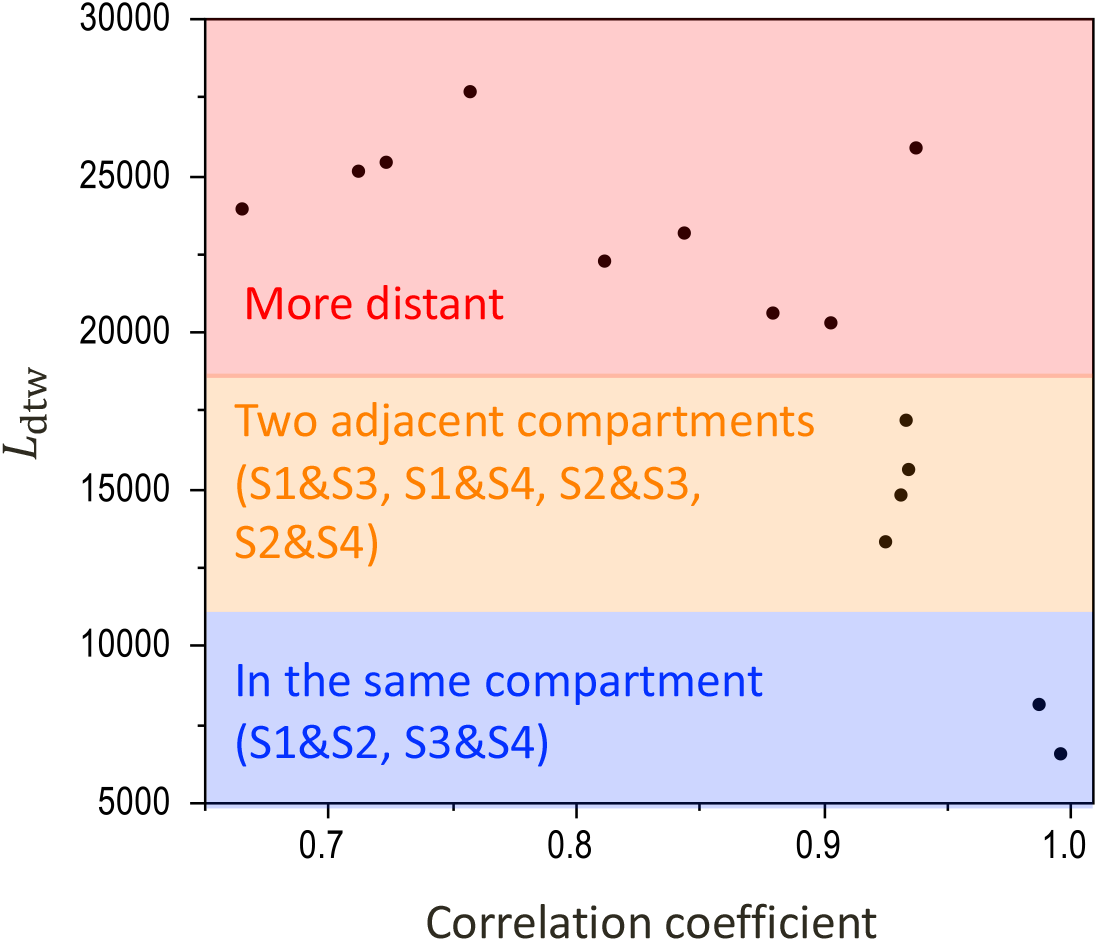
Scatter plot of correlation coefficient and minimum cumulative distance of each sensor data pair.

Fig. 7 shows the results of Large-eddy simulation (LES) for condition 1 using Flowsquare+ (https://fsp.norasci.com/), an open-sourced thermal fluid simulator. The simulation space is 32 (W) ⨯ 8 (D) ⨯ 4 (H) m in size, and each dimension is divided into 160 ⨯ 40 ⨯ 20 meshes. The fluid clay coefficient μ is 20 ⨯10^−6^, and the fluid density ρ is 1.2 kg/m^3^. The room temperature was set to 298 K, and exhaled air containing the virus at a temperature of 309 K was expelled from the two initially infected people at a ventilation linear velocity of 1 m/s. Figure 7 shows the concentration distribution of these exhaled breaths after 30 seconds. The infectious exhaled air stays in each compartment where the initially infected person is located. The computer simulation confirms that that the ventilation of the compartments is almost independent due to the partition effect of the PSS.

**Fig. 7.**
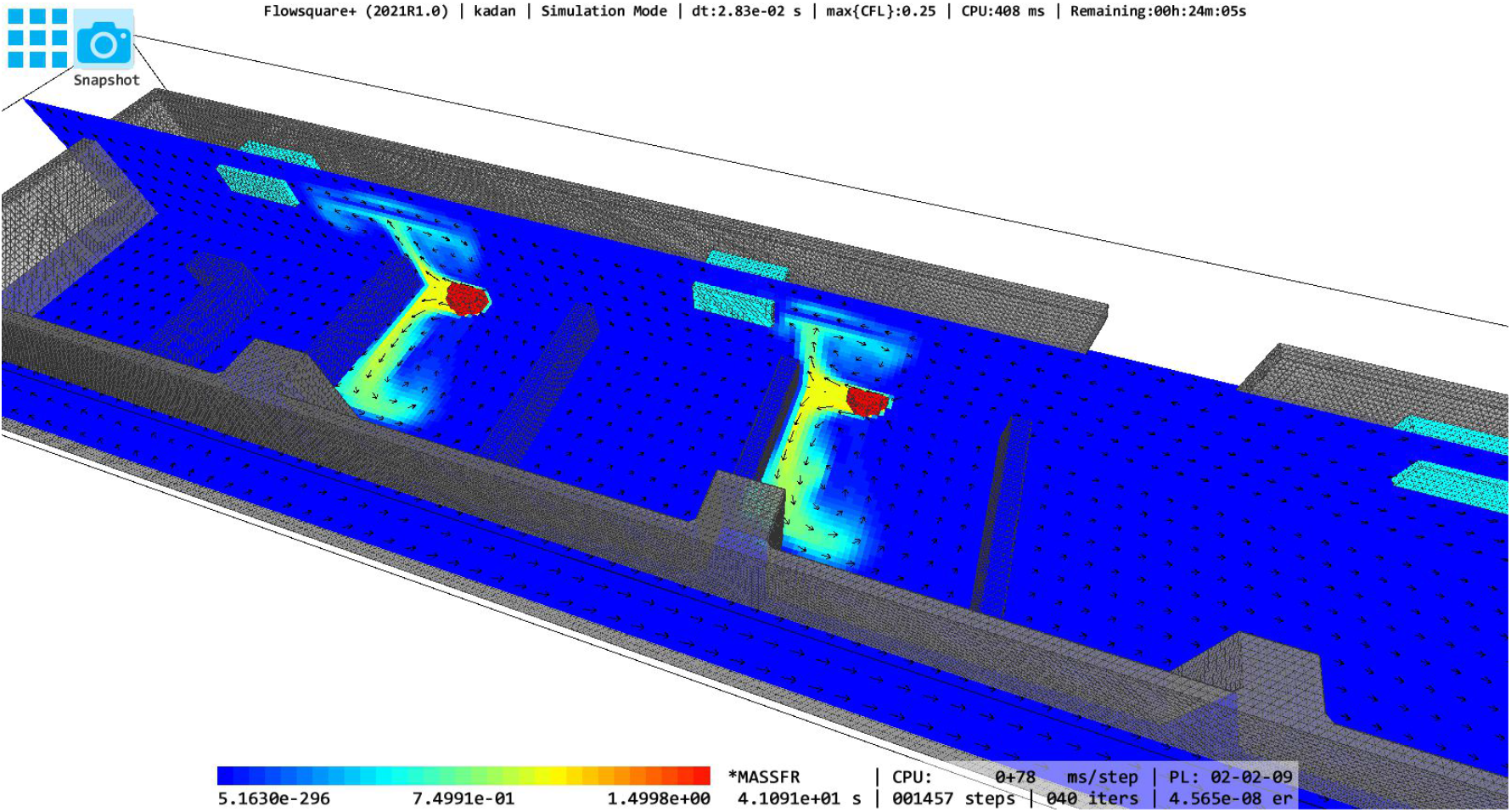
Concentration distribution of infectious aerosol at condition 1 reproduced by thermo-fluid simulation.

The air change rate of each compartment was estimated using the observed CO_2_-decreasing data and eq. (3). As shown in Fig. 8, the change in the normalized concentration of CO_2_ shows an approximately linear dependence on a single logarithmic chart, and the observations are plotted in the range of 95% confidence level of the model. The slope of the fitted line estimates the air change rate.

**Fig. 8.**
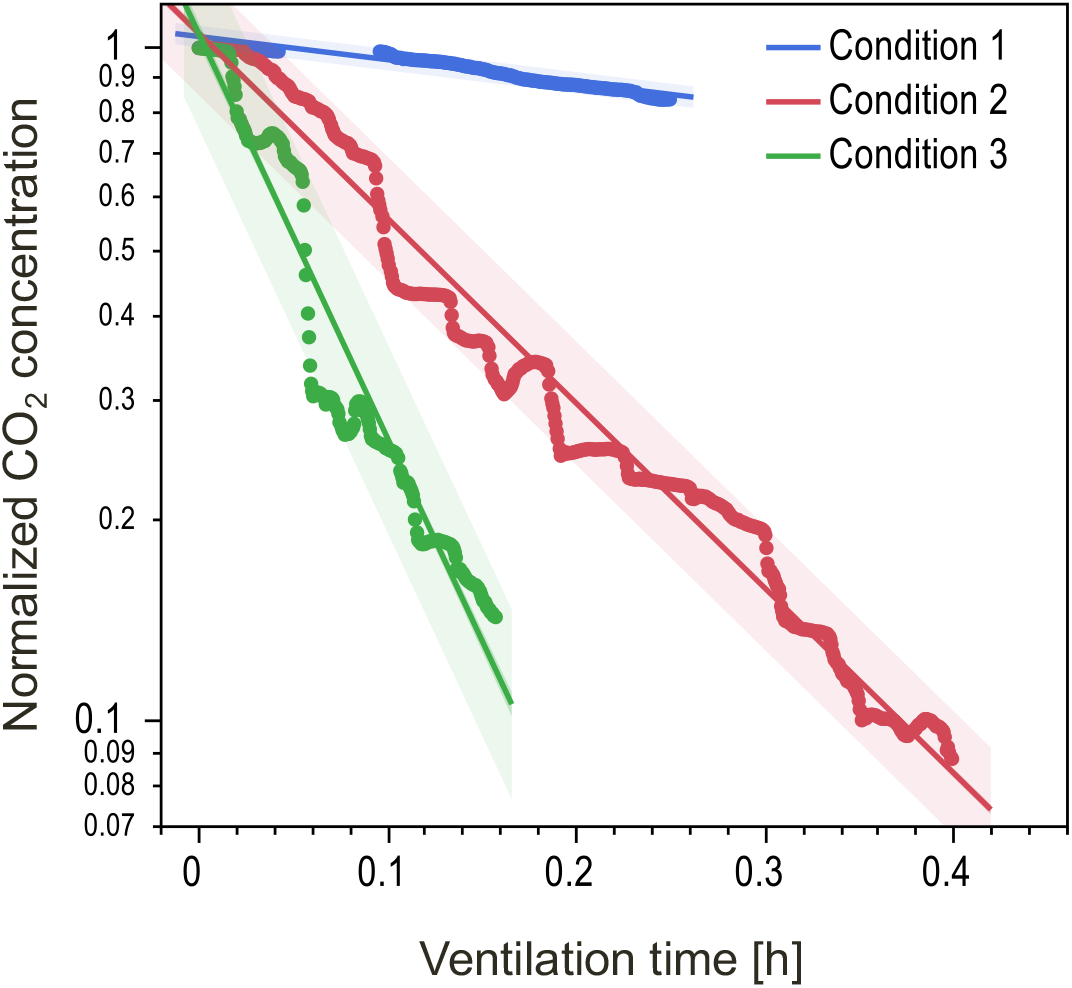
Time-series CO_2_ concentration plot as an example of local air change rate estimation based on eq. (3). Straight lines indicate least-square fitting and colored bands indicate 95% confidence interval of individual observations.

The comparisons of estimated air change rates of each condition are indicated in Fig. 9 with a 95% confidence interval. In condition 1, area A shows a shallow air change rate around 0.1 /h, implying poor ventilation. Area B shows an air change rate at around 0.8/h, which is significantly higher than that of condition 1. However, the value is not enough to prevent aerosol infection spread as reported for tuberculosis infection^18, 19^). Area D that is located near the entrance and area E, which is the largest, show an air change rate from 1.6 /h to 2.6 /h.

**Fig. 9.**
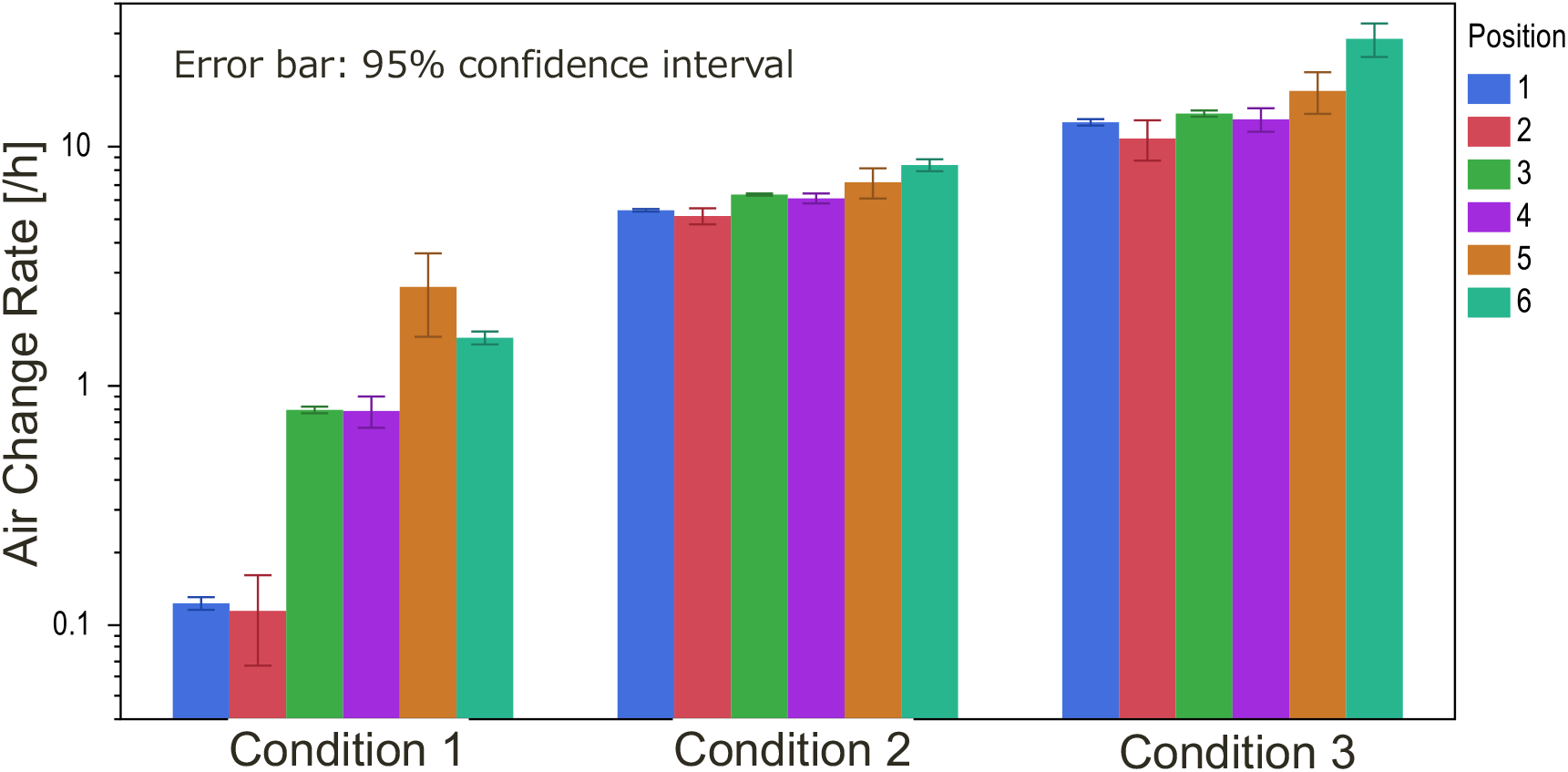
Bar plot showing the comparison of estimated air change rate of each location and condition.

In condition 2, the air change rate improves from 5.1 to 8.4 by ensuring ventilation routes through windows. Additionally, the difference in the air change rate is small, and it suggests opening windows uniformly improves the ventilation of each compartment. It is enhanced by the added fans as indicated by the air change rate value from 10 to 28 in condition 3.

Improvement in wind volume is correlated to the change in the air change rate, as shown in Fig. 10. The wind volume in condition 1 (Fig. 10(a)) is less as compared to those of conditions 2 and 3. The wind direction to windows is improved in condition 2 (Fig. 10(b)). The addition of fans in condition 3 enhances the airflow route (Fig. 10(c)), which seems to be related to the entrance as an air intake (see Fig. 1). After the decontamination operation of the COVID-19 outbreak, this office is operated with the ventilation of condition 3, and no additional PCR test positives were found for two months up to the time of this writing.

**Fig. 10.**
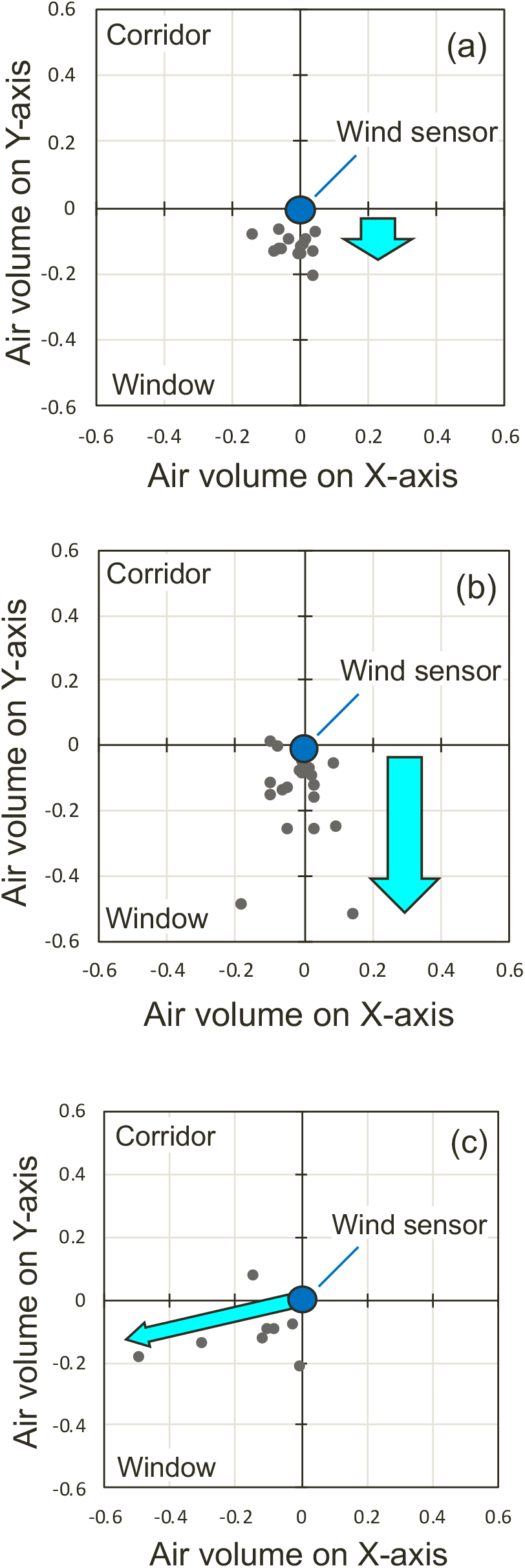
Scatter plot for the observation results of wind sensor for each condition. (a) Condition 1, (b) Condition 2, and (c) Condition 3.

## Conclusions

Statistical time-series data analysis of rising and fall patterns in the CO_2_ concentration indicates that PSSs separate the ventilation of compartments that contribute, prevent, and enhance the cluster of COVID-19. The computer simulation also confirmed that each compartment was ventilated almost independently due to the partitioning effect of the PSSs. Opening windows and using fans improve the ventilation in each compartment uniformly, and contribute to the prevention of cluster recurrence.

The installation of PSSs is supposed to prevent droplet infection and give users a sense of psychological security. However, if the ventilation capacity is reduced by PSSs, the risk of airborne infection by droplet nuclei must be considered. In addition, it is necessary to educate users so that they do not engage in risk-taking behaviors (e.g., removing masks, shouting) due to overconfidence in the effectiveness of PSSs.

This paper analyzed one case of a characteristic infection cluster in which PSSs may have been involved. As a next step, comprehensive simulations of how ventilation conditions change with the layout, height, and placement of ventilation equipment in PSSs would provide more general insight into the impact of PSSs.

## Data Availability

The data that support the findings of this study are available from the author, [Yo Ishigaki], upon reasonable request.

## Notes

The authors indicated no conflicts of interest.

## Acknowledgments

This work was supported by Research Grant Program of KDDI Foundation.

